# Duplication within 14q32.13 implicates a chimeric *CLMN*::*SYNE3* RNA transcript in cerebellar ataxia

**DOI:** 10.64898/2026.04.23.26350376

**Authors:** Thomas M Litster, Robert A Wilcox, Renée Carroll, Alison E Gardner, Nazzmer M Nazri, Cheryl A Shoubridge, Martin B Delatycki, Katja Lohmann, Marc Agzarian, Rafaela Turella Divani, Haloom Rafehi, Liam Scott, Gavin Monahan, Phillipa J Lamont, Catherine Ashton, Nigel G Laing, Gianina Ravenscroft, Melanie Bahlo, Eric Haan, Paul J Lockhart, Kathryn L Friend, Mark A Corbett, Jozef Gecz

## Abstract

The spinocerebellar ataxias (SCAs) are a clinically heterogenous group of neurodegenerative disorders that affect movement, vision, speech and balance.

Here, we reassign the linkage of SCA30 to 14q32.13 based on a cumulative LOD score >12. Within this interval we identified a 331 kb duplication, absent in population controls and not observed in >800 unrelated individuals with genetically unresolved cerebellar ataxia. RNA-Seq analysis of patient-derived lymphoblastoid cell lines revealed a splice-mediated chimeric transcript resulting from the duplication event. This transcript joined exon 1 of CLMN to exon 2 of SYNE3. In silico translation predicted that this chimeric transcript would produce a short N-terminal peptide corresponding to exon 1 of CLMN and the usually untranslated region of exon 2 of SYNE3 fused to the complete and in-frame SYNE3 protein. Transient overexpression of SYNE3 or the CLMN::SYNE3 fusion protein, in both HeLa cells and mouse primary cortical neurons, resulted in equivalent cellular outcomes including altered nuclear morphology and chromosomal DNA fragmentation. SYNE3 forms part of the linker of nucleoskeleton and cytoskeleton complex and is not usually expressed in cerebellar Purkyně neurons while, *CLMN* has a Purkyně specific expression pattern within the brain.

Our data suggests that ectopic expression of *SYNE3* in cerebellar Purkyně neurons, mediated by the *CLMN* promoter, leads to cerebellar atrophy and causes spinocerebellar ataxia in the SCA30 family. This is an example of Mendelian disease arising from a novel, chimeric transcript with a likely dominant negative effect. Chimeric transcripts are commonly associated with cancers, but they are not often associated with monogenic disorders. Detection of chimeric transcripts as part of structural variant analysis could increase the genetic diagnostic yield of Mendelian disorders.

## Introduction

Spinocerebellar ataxias (SCAs) are a genetically diverse group of neurodegenerative disorders that affect approximately 3 in 100,000 individuals.^1^ The core phenotype of SCAs comprises gait ataxia, nystagmus, and dysarthria.^2,3^ SCAs are caused by loss of cerebellar Purkyně neurons, which are inhibitory neurons that receive sensory and motor information, and transmit signals from the cerebellar cortex to control movement and coordination.^4,5^ Harding’s classification defines three clinical subtypes of SCAs, that are determined by their associated comorbidities:^6^ Autosomal Dominant Spinocerebellar Ataxia (ADCA) type I (core ataxia phenotype with additional pyramidal or extrapyramidal features, or amyotrophy), ADCA type II (core ataxia phenotype with pigmentary retinal degeneration), and ADCA type III (only core ataxia phenotype, known as pure cerebellar ataxia). This clinical heterogeneity is accompanied by considerable genetic heterogeneity, with over 40 genetically unique sub-types of SCA identified to date.^7^

The most frequent cause of SCAs are coding and non-coding short-tandem repeat (STR) expansions. STR expansions undergo both somatic and germline changes in the number, and sometimes base composition, of the repeated units. The dynamic nature of STR expansions is associated with anticipation where symptoms onset earlier and are more severe in successive generations. Multiple pathogenic splice-altering, or protein-damaging, single nucleotide variants (SNVs) and small indels have been implicated in SCA though collectively they account for a small proportion of individuals living with SCA.^7^ Structural variants (SVs) are alterations to large segments of the genome and include copy number variants (CNVs), translocations, inversions and novel sequence insertions. SVs frequently result in altered expression of the genes included within, and adjacent to the variant, which can be detected by RNA-seq.^8^ SVs and CNVs are rarely implicated in SCAs with recurrent whole-gene deletions of *ITPR1* or *FGF14,* a single family with an intragenic deletion in *SPTBN2*, a 260 kb duplication implicated in SCA20 and a 7.5 Mb duplication implicated in SCA39 identified to date.^9–16^

The genetic diagnostic rate for hereditary ataxias using genome sequencing and analyses of STR, SNV, indels and SVs was 33-38%, while with exome sequencing a similar approach yielded 24%.^17–19^ Here, we aimed to identify the genetic cause of SCA (ADCA type III) in a multigenerational family of European ancestry using a combination of linkage, genome sequencing and RNA-seq analyses.

## Materials and methods

### Cell Culture

Patient derived Epstein-Barr virus immortalised lymphoblastoid cell lines (LCLs) were cultured in RPMI 1640, GlutaMAX-l and 25 mM HEPES (Gibco: 72400-047).

HeLa cells were cultured in DMEM, GlutaMAX-l, 5.7 g L^-1^ D-Glucose and 110 mg L^-1^ Sodium Pyruvate (Gibco: 10669-010). HeLa and Cos7 cell lines were split 1:15 when at 80%-90% confluency using 1X Trypsin (Sigma: 59417C). All media was supplemented with 10% foetal bovine serum (FBS) and 1% Penicillin-Streptomycin (100 U mL^-1^ penicillin, 100 μg mL^-1^ streptomycin), (Gibco: 15140-122).

All cells were cultured in an environment of 5% CO_2_ at 37 °C.

### DNA extraction

DNA was extracted from LCL pellets using the DNeasy Blood & Tissue Kit (Qiagen: 69506) following manufacturer’s instructions. DNA was extracted from whole blood using the Maxwell RSC Whole Blood DNA Kit (Promega: AS1520). Purity of extracted DNA was assessed using NanoDrop spectrophotometry and quantity was assessed using Qubit dsDNA Broad Range Quantification Assay (Invitrogen: Q32835). Quality of whole blood gDNA was assessed with agarose gel electrophoresis. Quality of LCL gDNA was assessed with TapeStation, using the Genomic DNA ScreenTape (Agilent: 5067-5365) and Genomic DNA Reagents (Agilent: 5067-5366).

### RNA extraction

Total RNA was extracted from LCL pellets using a QIAshredder (Qiagen: 79654) to homogenise, and RNeasy mini kit (Qiagen: 74106) to purify RNA, following manufacturer’s instructions. RNA purity and concentration was assessed with NanoDrop spectrophotometry and quality was assessed by agarose gel electrophoresis.

### Genome-wide parametric linkage analysis

Whole blood gDNA from 42 family members (8 affected males, 7 unaffected males, 19 affected females and 9 unaffected females) was genotyped with Illumina Global Screening Array (GSA) v3 BeadChips as a service by the Australian Genome Research Facility. Individuals were recruited between 1993 to 2023, based on self-reported family relationships and their informed consent for genetics research. Final reports were generated by Illumina GenomeStudio (v2.0.4) using the “GSA-24v3-0_A2” manifest and “GSA-24v0_AI_ClusterFile” cluster files.

Due to the size of the SCA30 pedigree, it was divided into four non-overlapping sub-pedigrees, each of which underwent separate linkage analyses (Supplemental Fig. 1). To make these analyses comparable, LinkDataGen^20^ was used to select 11,474 SNPs using the complete SCA30 pedigree as input matched to the CEU population in the “annot1m.txt” annotation file from Illumina. To ensure these SNPs were used for linkage analysis of all sub-pedigrees, LinkDataGen bin size was set to 0, a list of the selected SNPs was provided with the-fileKeepSNPs option, and all other SNPs were excluded using the -fileRemoveSnps option. MERLIN v1.1.2^21^ was used to run the linkage analysis with an autosomal dominant disease model (disease allele frequency of 0.001, disease penetrance of 0%, 99% and 99% for 0/0, 0/1 and 1/1 genotypes, respectively). The LOD scores from loci where all four sub-pedigrees had a positive LOD score were summed to combine the results from each of the four linkage iterations.

### Short-read genome sequencing analysis

Whole blood gDNA extracted from three affected females (V.47, V.56, V.104) was sent to NovoGene Hong Kong (for V.47 and V.56) for sequencing on the Illumina NovaSeqX Plus platform or to Kinghorn Centre for Clinical Genomics (for V.104) for sequencing on Illumina HiSeq X Ten platform.

Short-read GS data was mapped to the hs38DH human reference genome build with Burrows-Wheeler aligner (BWA-MEM) v0.7.17q using default settings for alignment a genome with alt contigs.^22^ Variants were jointly called in combination with GS from 21 additional unrelated individuals using genome analysis toolkit (GATK) v4.2.6.1 haplotype caller, following best practices^23^ and annotated with ANNOVAR.^24^ Non-coding variants were annotated with CADD^25^ v1.6, ReMM^26^, and ENSEMBL Variant Effect Predictor^27^ to assess their functional impact.

Variants with a frequency greater than 1/5000 (0.0002) minor allele fraction in gnomAD v2.1.1 and variants with an allele fraction above 3/24 (0.125), within the group of 24 genomes from which variants were jointly called were removed to retain only variants private to the SCA30 family.

Expansion Hunter^28^ and Expansion Hunter DeNovo^29^ v0.9.0 were used to scan the genome for short tandem repeat expansions. Structural variants were called using GRIDSS2^30^ and Parliament2^31^ and annotated with AnnotSV.^32^

### Oxford Nanopore long-read DNA sequencing

Genomic DNA extracted from blood from IV.104 or LCL from V.16 was prepared using the SQK-LSK109 ligation sequencing kit (Oxford Nanopore Technologies), following the manufacturer’s protocol and sequenced on a R9.4.1 flow cell using a PromethION 48 device as a service by NovoGene.

FAST5 files were basecalled with bonito v0.6.2 (Oxford Nanopore Technologies) with the dna_r9.4.1_e8_sup@v3.3 basecalling model and the resulting fastq files were filtered to remove any reads with an average quality score below 7 with NanoFilt v2.8.0.^33^ Minimap2^34^ v2.17 was used to align filtered fastq to human refence build GRCh38 (GCA_00000140.15) with alt contigs removed. Structural variants were called with Sniffles2^35^ v2.0.7 in population calling mode along with six additional ONT long-read genomes then annotated with AnnotSV.^32^

### Chromosome 14 duplication PCR

The novel breakpoint produced by the chromosome 14 duplication, and a region flanking the duplication, was amplified by duplex PCR using primers chr14_dup_F1 at 40 nM, chr14_dup_F2 and chr14_dup_R1 at 400 nM (Supplemental Fig. 2, Table S1). PCRs comprised of all three primers, 50 ng template DNA, 0.4 mM each dNTP and 1U Taq DNA polymerase in 1 x PCR buffer (Sigma: 11418432001) were run with the following conditions: one cycle of 94 °C for 3 minutes, 35 cycles of 94 °C for 30 seconds, 61 °C for 30 seconds, and 72 °C for 60 seconds, followed by 3 minutes at 72 °C.

### Short-read RNA-sequencing analysis

Total RNA, extracted from LCLs of three affected females (V.56, V.61, VI.16), an affected male (V.82) and four unrelated and unaffected females was sent to the South Australian Genomics Centre (Adelaide) for ribosomal RNA depletion and library preparation. Stranded libraries were prepared using the Nugen Universal Plus Total RNA seq protocol, followed by the MGIEasy Universal Library Conversion Kit. Libraries were sequenced with the MGI DNBSEQ-G400 device to generate a minimum of 1x10^8^ 98bp paired-end, strand-specific reads per sample.

RNA-seq data was aligned to the GRCh38 human genome reference build (grch38_tran available from https://genome-idx.s3.amazonaws.com/hisat/grch38_tran.tar.gz) using HISAT2.^18^ Alignments were provided to DROP^36^ v1.3.2, along with a cohort of 107 other unrelated RNA-seq samples for detection of outlier expression events, and 30 for outlier splicing events.

For differential expression analyses, transcripts matching ENSEMBL v84 annotations were quantified using Salmon v 1.10.0. Comparisons between affected individuals in the SCA30 family and controls were made with a moderated likelihood ratio test from the edgeR package in Bioconductor.^37^ Genes with false discovery rate (FDR) < 0.05 were classed as differentially expressed.

### Cloning expression vectors

The mEmerald-Nesprin3-C-18 plasmid (referred to here as mEmerald-SYNE3) was a gift from Michael Davidson (Addgene plasmid # 54203; http://n2t.net/addgene:54203; RRID: Addgene_54203).

To create the mEmerald-CLMN-SYNE3-Fusion expression vector, the *CLMN*::*SYNE3* chimeric transcript from polyA+ cDNA generated from an affected individual was amplified using primers CLMN_SYNE3_cloning_F and CLMN_SYNE3_cloning_R (Supplemental Table S1) and Phusion High-Fidelity DNA Polymerase as per the manufacturer’s instructions (Thermo Fisher Scientific: M0530S). The PCR cycling conditions were: one cycle of 98 °C for 3 minutes, 40 cycles of 98 °C for 15 seconds, 59 °C for 60 seconds, and 72 °C for 60 seconds, followed by 10 minutes at 72°C. The *CLMN*::*SYNE3* chimeric transcript PCR product digested with *Xho*I (NEB: R0146) and *Bss*HII (NEB: R0199) was cloned into the corresponding sites in the mEmerald-SYNE3 vector.

To create the mEmerald-Empty-Vector, the mEmerald-Nesprin3-C-18 was digested with *Xho*I (NEB: R0146) and *Kpn*I-HF (NEB: R3142S), blunt ended using T4 DNA Polymerase (NEB: M020) and then re-ligated with the T4 DNA ligase (NEB: M0202), following manufacturer’s instructions.

### Preparing plasmids for transfection

Vectors transformed into DH5α high efficiency chemically competent *E. coli* (NEB: C2987H), were grown overnight at 37 °C in a shaking incubator in 200 mL LB media supplemented with 50 μg mL^-1^ kanamycin.

All vectors were purified using the GeneJet Endo-free Plasmid Maxiprep Kit (Thermo Fisher Scientific: K0861), following manufacturer’s instructions.

### Transfection of HeLa and Cos7 Cells

HeLa cells were plated on poly-L-lysine coated coverslips and Cos7 cells on uncoated coverslips 24 hours prior to transfection at a density of 5 x 10^4^ cells per well in a 12 well plate. Cells were transfected with 1 µg of either the mEmerald-Empty-Vector, mEmerald-SYNE3, or mEmerald-CLMN-SYNE3-Fusion vector using Lipofectamine 3000 (Invitrogen: L3000015), following manufacturer’s instructions. Media was exchanged at 4 hours post-transfection. Cells were fixed in 4% paraformaldehyde for 15 minutes at 24 hours post transfection.

### Establishment of mouse primary cortical neuronal cultures

Cortical neurons were isolated from embryonic day (E) 17.5 C57BL/6 mice as previously described.^38^ In brief, neuronal suspensions were plated at 5 x 10^5^ cells/well on 0.1% poly(ethyleneimine) solution (Merck: P3143,) and 20 μg mL^-1^ laminin (Merck: L2020-1mg) coated coverslips in 24-well plates (Sigma: CLS3527,). Neurons were initially incubated in neural seed media (Neurobasal-A media (Gibco: 12348017), 10% Fetal Bovine Serum (FBS) (Gibco: 10099141), 2% B-27 (Gibco: 17504044), 1% Penicillin-Streptomycin and 1% L-Glutamine (Gibco: 25030081) immediately after plating and cultured with 5% CO_2_ at 37 °C. Media was exchanged to neural feed media (neural seed media without FBS) supplemented with 1% L-glutamic acid monosodium (MSG), (Sigma: G5889-100G) at 24 hours post plating. Half media changes with neural feed media were completed every two days.

### Transfection of Primary Mouse Neurons

Primary cortical mouse neurons were transfected at *in vitro* day 4. Neurons were transfected with 1 µg of either the mEmerald-Empty-Vector, mEmerald-SYNE3, or mEmerald-CLMN-SYNE3-Fusion vector using Lipofectamine 2000 (Invitrogen: 11668027), following manufacturer’s instructions, but increasing the volume of Lipofectamine 2000 to 4 μL per well. At 4 hours post-transfection, cells were washed with Neurobasal-A media (Gibco: 12348017), before adding neural feed media. Four coverslips were transfected per expression vector with two fixed at 24 hours post-transfection (*in* vitro day 5) and two fixed 72 hours post transfection (*in vitro* day 7). Neurons were fixed with 4% paraformaldehyde for 15 minutes.

### Immunofluorescence

Fixed cells were permeabilised and blocked in a solution of PBS (Sigma: P4417), 0.1% Tween20 (Sigma, P1379), 5% horse serum (Gibco: 16050-130) for 1 hour. Cells were incubated overnight with primary antibodies in a solution of PBS, 0.1% Tween20, 0.5% horse serum at 4°C. HeLa cells were incubated with either anti-Plectin or anti-Vimentin primary antibodies (Table S2). Primary mouse neurons were incubated with anti-Plectin and anti-β-III Tubulin primary anitbodies (Table S2). Cells were washed 8 times with PBS, 0.1% Tween20 (PBST) and then incubated for 1 hour at room temperature with secondary antibodies (Table S3), followed by 10 washes with PBST. Cells were mounted with ProLong Diamond Antifade Mountant with DAPI (Invitrogen: P36962).

Cells were imaged with a Zeiss Axio Imager M2 and Zeiss Axio Scan.Z1 Slide Scanner. When imaging with the slide scanner, images of HeLa cells were taken over a single plane of focus at 20x magnification. Images of primary mouse neurons were taken across four planes of focus spanning 10 μm at a magnification of 43x.

### Immunofluorescence image analysis

Nuclear area was analysed from micrographs with ImageJ v2.14/1.54f using Gaussian blur (sigma = 3) and auto-thresholding with Huang Dark model. A mask was created, on which watershed was run to split up overlapping nuclei. Measurements of nuclear area were made from this mask, excluding any regions of interest (ROIs) overlapping with the edge of the image, an area smaller than 10 pixels^2^, or with a circularity lower than 80. These ROIs were then overlayed onto images of the green channel of the same image, and measurements of signal intensity were taken to identify transfected cells.

Neuronal nuclei were counted using Zeiss Zen software v3.12 from multichannel images of transfected primary mouse neurons. The red channel (β-III Tubulin) was used to identify neuronal nuclei. The green signal (mEmerald) was used to identify transfected neurons and classified as either perinuclear or cytoplasmic. The blue channel was used to classify nuclei of transfected neurons as either normal, small round, or fragmented. A target of classifying 300 nuclei across two replicates for each time point and transfection condition was set.

## Ethics statement

Research involving human participants was carried out with approval from the Women’s and Children’s Health Network research ethics committee, HREC2361/3/2026, The Royal Children’s Hospital Human Research Ethics Committee (HREC #28097). Informed consent was obtained from participants in this study. Harvest and culture of mouse primary neurons was approved by the University of Adelaide Medical Animal Ethics Committee; M-2023-017.

## Data availability

GS and RNA-seq data generated in this study is available via request to the corresponding author and is subject to approval for use by the Women’s and Children’s Health Network human research ethics committee.

Mouse single nuclei data^39^ is available from https://docs.braincelldata.org/downloads/index.html

Human developmental bulk RNA seq data from EvoDevo^40^ is available from https://www.ebi.ac.uk/biostudies/arrayexpress/studies/E-MTAB-6814

## Results

We identified a multigenerational family of European origin (Fig. 1A) in which affected members were later identified as directly related to the SCA30 family (OMIM: 613371).^41^ Symptoms in this family typically began in the 5^th^ to 6^th^ decade of life, with the earliest known onset in the 4^th^ decade. Affected individuals initially presented with staccato speech, with midline balance impacted 5 to 10 years after onset. As disease progressed, appendicular coordination was impacted. Some affected individuals presented with nystagmus, however no other visual deficits were observed. No cognitive or memory deficits were present, there was no evidence of genetic anticipation, and no pyramidal features were observed. Imaging showed severe, symmetrical, bilateral cerebellar hemisphere atrophy, including the superior and dorsal cerebellar vermis (Fig. 1B). No involvement of brainstem or supratentorial structures was seen.

**Figure 1:**
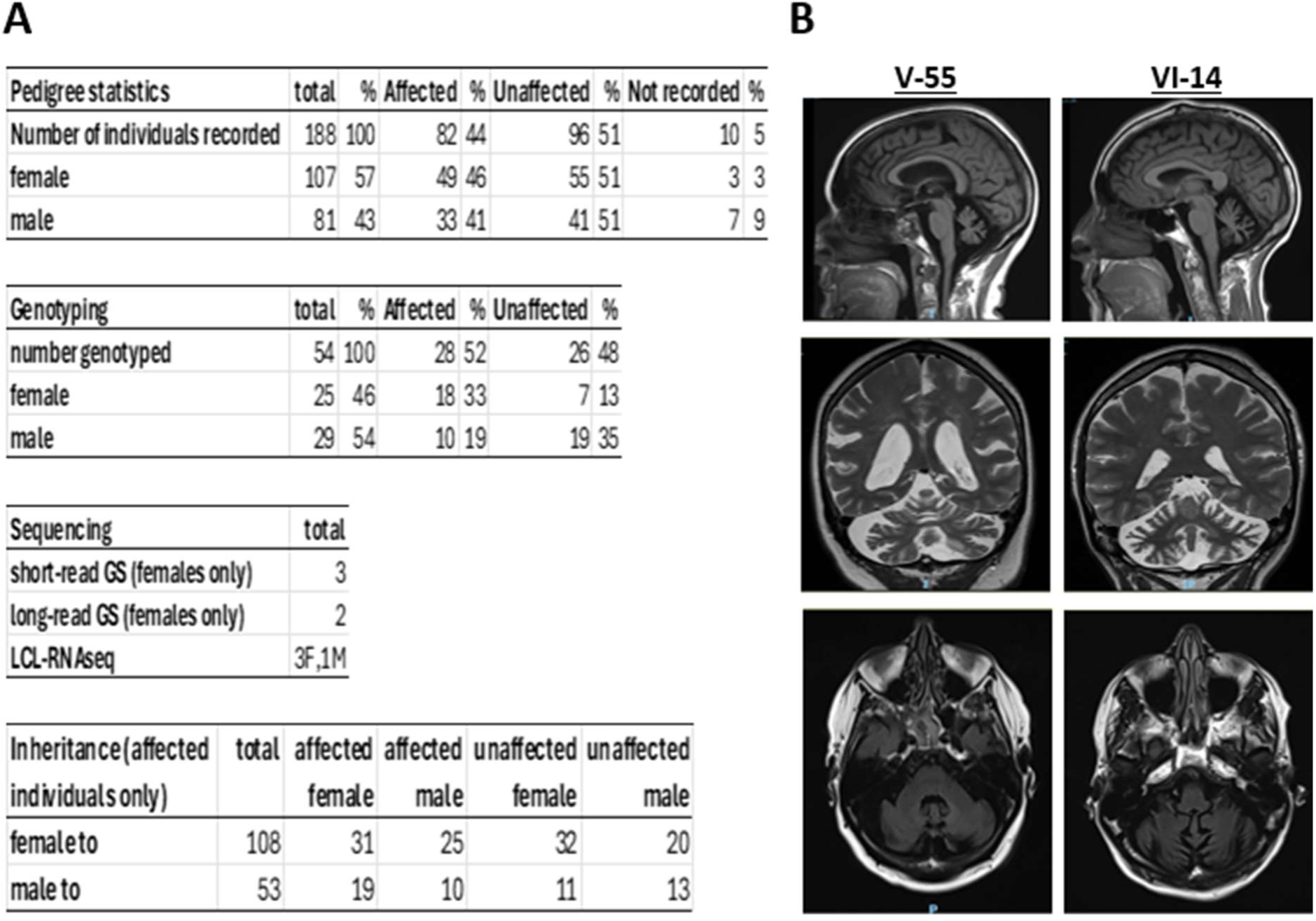
SCA30 is an autosomal dominant ataxia characterised by distinct cerebellar atrophy on brain MRI. (A) Tables describe the details of a six-generation pedigree with an autosomal dominant inheritance pattern of pure cerebellar ataxia. F; female, M; male, GS; genome sequencing (B) T1 fluid attenuated inversion recovery (FLAIR) turbo spin echo (TSE) sagittal (top row), T2 TSE coronal (middle row) and T2 FLAIR TSE axial (bottom row), MRI scans at 3 Tesla of individuals V-55 and VI-14 showing severe, symmetrical, bilateral cerebellar hemisphere atrophy, including atrophy of the cerebellar vermis. There is no atrophy of the supratentorial brain, midbrain, pons or medulla.

When initially reported, SCA30 linkage was established to 4p34.3-q35.1.^41^ We performed genome-wide parametric linkage analysis using SNP genotype data from 46 individuals of the SCA30 family. These individuals were split into four non-overlapping sub-pedigrees (Supplemental Fig. 1). Genome wide linkage analysis revealed no significant interval on chromosome 4, however a single significant linkage peak on chromosome 14 was detected in all four sub-pedigrees (Supplemental Fig. 1). The maximum LOD scores for each of the sub-pedigrees were 2.41, 3.14, 3.73, and 3.01, respectively. Summing loci where all sub-pedigrees had a positive LOD score refined this interval to 14q32.13 between SNP markers rs11621961 and rs4900285 (GRCh38: chr14:94303139-95628369). The maximum combined LOD score was 12.28 (Fig. 2A). The 1.3 Mb interval contained 12 protein coding genes (*SERPINA6*, *SERPINA1*, *SERPINA12*, *SERPINA9*, *SERPINA3*, *SERPINA4*, *SERPINA5*, *GSC*, *DICER1*, *CLMN*, *SYNE3*, *GLRX5*). No genes or mapped loci previously implicated in SCA were located within the linkage interval. The closest known SCA gene is *ATXN3* (SCA3 / Machado-Joseph disease, OMIM: 109150), which is located 3 Mb centromeric to the linkage interval.^42^

**Figure 2:**
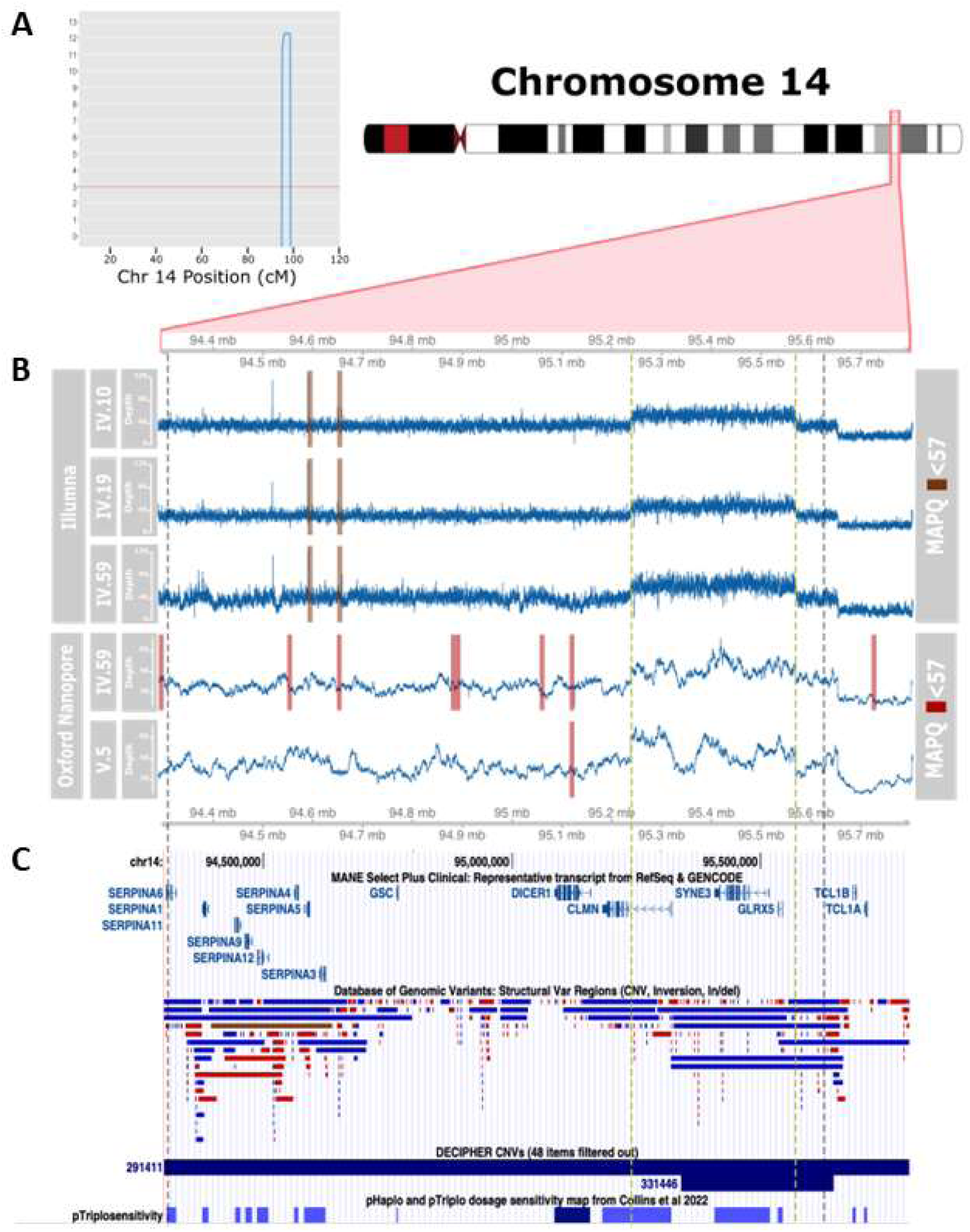
Genome-wide parametric linkage to an interval on 14q32.13, which contains 13 protein coding genes. (A) Plot of cumulative positive LOD scores from each of the four sub-pedigrees. A single interval with boundary markers rs11621961-rs4900285 corresponding to chr14:94303139-95628369 (GRCh38), with a maximum LOD score of 12.28 is shown. (B) Summary of depth of coverage and mapping quality of genome sequencing generated with Illumina and Oxford Nanopore Technologies (ONT) for the chromosome 14 linkage interval. Average coverage (blue) was calculated in 100 bp windows. Average mapping quality was calculated in 1 kb windows, with regions highlighted in red (Illumina) or brown (ONT) having an average mapping quality below 57. Grey dashed lines running down the figure mark the boundary of the linkage interval, and yellow dashed lines mark the boundary of the chromosome 14 duplication. (C) The chromosome 14 linkage interval encompasses 13 protein coding genes (GENCODE V44), with the chromosome 14 duplication affecting three genes. Population level structural variants from Database of Genomic Variants show some copy-gain variants with similar boundaries to the SCA30 duplication, however the breakpoints of these variants do not match the chromosome 14 duplication. DECIPHER CNVs shows pathogenic copy gain (blue) and copy loss (red) variants affecting the same regions as the SCA30 duplication, note that no variants match the boundaries of the SCA30 duplication. pTriplosensitivity values of genes in the linkage interval show that only DICER1 is sensitive to duplication (dark blue).

To identify the genetic cause of disease in the SCA30 family we initially analysed short-read genome sequencing (GS) from affected individuals V.47, V.56 and V.104. There were no regions within the linkage interval where all genomes had less than 15-fold depth of sequencing coverage. There were two 1 kb windows that had mapping quality below 57; a previously used cutoff indicating an inability to interrogate that region for causative variants,^43^ however, these did not overlap with known genes (Figure 2B and 2C). Two SNVs, chr14:g.94353020A>T and chr14:g.95545106G>A (rs9934694), that passed our population filtering criteria were identified within the linkage interval of all affected individuals. Neither of these occurred within a gene or were predicted to be damaging (Supplemental Table S4). No rare and private variants were detected in known SCA genes (https://neuromuscular.wustl.edu/ataxia/domatax.html; accessed 1^st^ December 2025), and no known pathogenic variants implicated in SCA from the ClinVar Database (accessed 21^st^ of October 2025) were present in the individuals we sequenced. Given the enrichment of causal repeat expansions in SCAs, the GS data was scanned for known and novel STR expansions however no repeat expansions previously implicated in SCA nor novel expansions within the linkage interval were detected.

To identify potentially causative structural variants, we generated Oxford Nanopore Technologies (ONT) long-read GS data for two affected individuals from the SCA30 family (V.104 and V.16). Structural variants were called from the ONT GS and from the short-read GS. Two rare shared structural variants were detected in both long and short read GS data that were unique to the affected individuals in the SCA30 family. These were a 331 kb duplication (NC_000014.9:g.95239046_95570197dup) which was within the linkage interval (Fig. 2B, Table S5), and a 148 kb deletion (NC_000014.9:g.95654685_95802922del) which was outside of the linkage interval and thus unlikely to be causative. The 5’ boundary of the novel 331 kb duplication occurred 80 kb within intron 1 of *CLMN* and the structural variant completely duplicated coding genes *SYNE3* and *GLRX5*, along with a non-coding gene, *SNHG10* (Fig. 2C). Using PCR to amplify across the unique junction within the tandem duplication (Supplemental Fig. 2), this variant segregated with all tested affected individuals and was also detected in five unaffected males (IV.33, IV.34, V.42, V.63 and V.105) in the SCA30 family (Table 1). This PCR was used to screen 436 individuals with ataxia that are currently unresolved for genetic causes from a matched population. This identified one individual with the NC_000014.9:g.95239053_95570195dup variant who had no known connection to the SCA30 family and for whom further information could not be obtained. Additionally, no individuals from a cohort of 452 individuals from Northern Germany with genetically undiagnosed ataxia had this duplication. A cohort of 268 anonymous controls sourced from the Australian Red Cross blood bank were also screened, all of whom were negative for the duplication.

While no duplications identical to NC_000014.9:g.95239053_95570195dup were identified in population genetic databases, four overlapping duplications were found (Fig. 2C). One ultra-rare duplication was identified in the gnomAD^17^ v4.1 SV callset (DUP_14_39057; NC_000014.9:g.95320062_95667131dup) at a frequency of 2/126,080. Additionally, three similar duplications were identified in the Database of Genomic Variants (DGV) (v107):^44^ dgv89e55; NC_000014.9:g.chr14:95292222_95675258dup, nsv565631; NC_000014.9:g.chr14:95320320_95666158dup, and nsv1042858; NC_000014.9:g.chr14:95326385_95660745dup (Fig. 2C). One individual with a NC_000014.9:g.95246699_95781052x3 triplication was present in DECIPHER v11.34 (DECIPHER ID: 2922343).^45^ The phenotype of this individual did not match SCA30 and they had no recorded family history of SCA. In each case, the locations of the breakpoints in these previously identified duplications or triplications were not identical to the NC_000014.9:g.95239046_95570197dup in the SCA30 14q32.13 linkage interval.

To investigate the effect of the NC_000014.9:g.95239046_95570197dup duplication on gene expression, we performed RNA-seq on total RNA extracted from lymphoblastoid cell lines (LCLs) of three affected females (V.56, V.61 and VI.16) and an affected male (V.82) from the SCA30 family compared to four unrelated female controls. We observed RNA-seq reads mapped to the reverse strand of chromosome 14 in the intergenic space between *GLRX5* and the 3’ boundary of the duplication in affected individuals but not controls (Fig. 3A). This suggested that the promoter of the partially duplicated copy of the reverse strand *CLMN* gene was transcriptionally active. Read pairs with larger than expected and negative insert size were identified in affected individuals between exon 1 of *CLMN* and exon 2 of *SYNE3*, suggesting the formation of a *CLMN::SYNE3* splice-mediated chimeric transcript (Fig. 3B). The existence of this chimeric transcript was confirmed by ONT sequencing of RT-PCR products from a primer pair complementary to sequence within exon one of *CLMN* and exon two of *SYNE3* in cDNA derived from LCLs of four affected individuals from the SCA30 family (Supplemental Fig. 3). The *CLMN*::*SYNE3* chimeric transcript was not seen in unrelated controls. *In silico* translation of the *CLMN*::*SYNE3* chimeric RNA predicted exon one of *CLMN* and the first 14 bases (which are normally untranslated) of exon two of *SYNE3*, would add a short N-terminal peptide of 32 residues, with no encoded functional domain, in-frame to the full-length open reading frame of SYNE3 (Fig. 3B). Comparing the whole transcriptome of affected individuals to controls identified 31 differentially expressed genes (FDR < 0.05) and of these, *GLRX5* was the only gene within, or adjacent to, the chromosome 14 duplication that was significantly upregulated (Fig. 3C, Supplemental Table S6). ShinyGO^46^ v0.82 was used to perform gene ontology enrichment analysis on the list of all differentially expressed genes, using all genes expressed in LCL as the background and identified potential, though relatively minor, impacts on cell metabolism, mRNA export, and histone demethylases (Supplemental Table S7). Of the genes contained within, or adjacent to, the chromosome 14 duplication, only *DICER1* is sensitive to an increase in copy number,^47^ but its expression was not altered in LCLs (Fig. 3C). Additionally, no outlier expression or splicing events that were shared by the affected individuals were detected. As such, the *CLMN*::*SYNE3* chimeric transcript was targeted for further investigation as the potential cause of the cerebellar ataxia in the SCA30 family.

**Figure 3:**
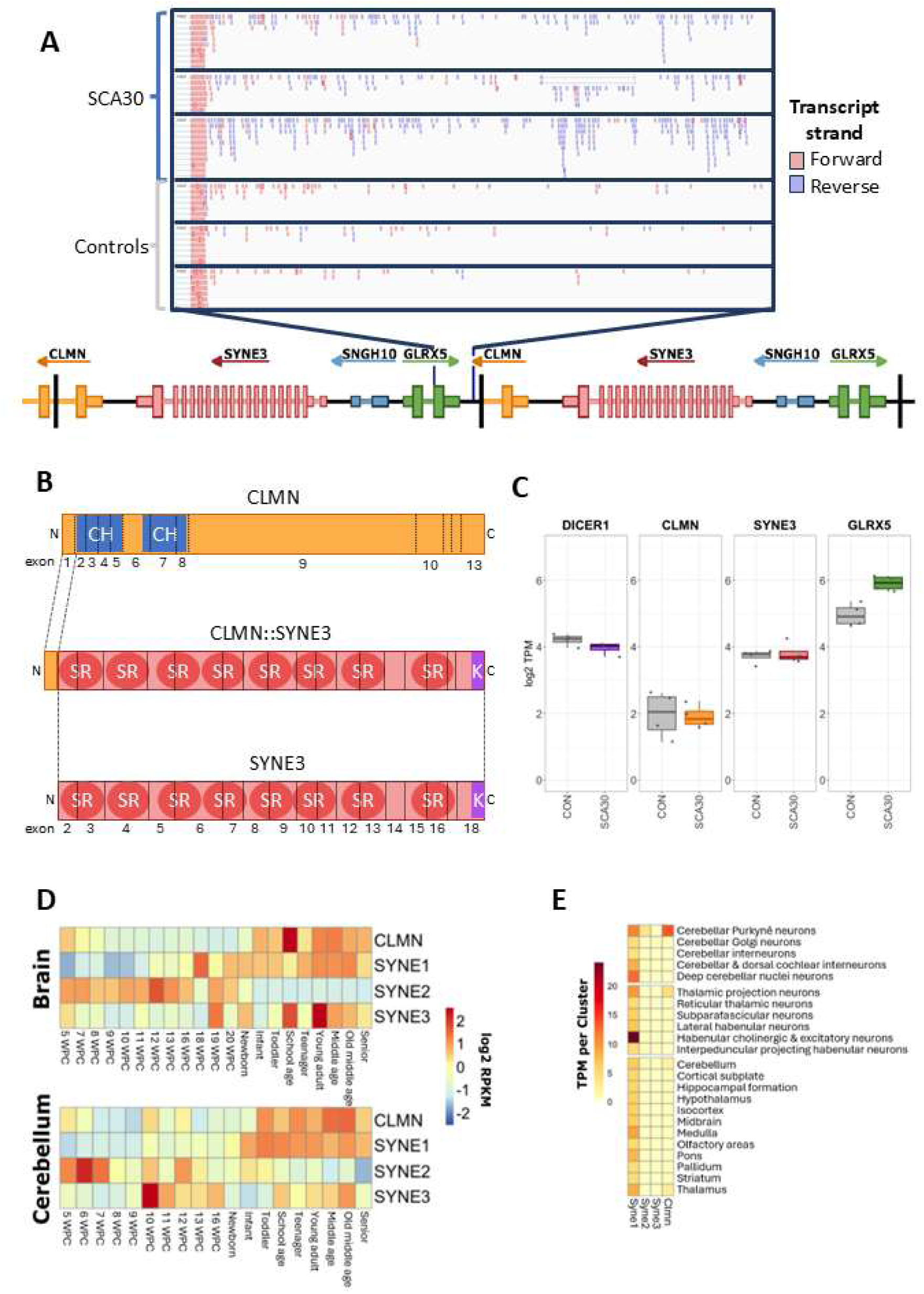
The chromosome 14 duplication gives rise to a splice-mediated chimeric fusion transcript between CLMN and SYNE3. (A) An IGV screen shot of LCL RNA-seq data shows an example of transcription (reads in blue) on the reverse strand of chromosome 14 in the intergenic space commencing from the 3’ breakpoint of the NC_000014.9:g.95239046_95570197dup variant downstream from GLRX5 three individuals from SCA30 that was not detected in any unrelated control. (B) Diagrammatic representation of SYNE3, CLMN and the CLMN::SYNE3 proteins overlaid with their respective exon structure. CLMN is a 1003 aa protein with two calponin homology domains (CH) indicated in blue. SYNE3 encodes a 976 aa protein with 8 spectrin-like repeats (SR) indicated with the darker pink circles and a C-terminal KASH domain (K) indicated in purple. The CLMN::SYNE3 protein is a fusion of exon 1 of CLMN to exon 2 of SYNE3 as shown. The amino and carboxyl termini are indicated with N and C respectively. (C) Differential gene expression analysis shows GLRX5 expression was upregulated in SCA30 samples (Log2 fold change 0.99, FDR=0.0039). CLMN, SYNE3, and DICER1, which closely neighbours the SCA30 duplication, showed no significant changes to their expression between SCA30 family members and unrelated controls. (D) Heat map showing expression of SYNE1, 2, and 3, and CLMN, across multiple developmental and post-natal time points in the human brain. The cerebellum shows an increase in CLMN expression with time, and a reduction in SYNE3 expression in adults. Weeks post-coitum (WPC). (E) Heatmap showing gene expression of Clmn, Syne1, 2, and 3, as average counts per nucleus per cluster, in mouse brain snRNAseq data. Syne1 has a general neuronal expression pattern, whereas Syne2 and Syne3 show a non-neuronal pattern. Clmn is expressed in thalamic projection neurons in the thalamus and Purkyně neurons in the cerebellum.

Synaptic Nuclear Envelope Protein 3 (SYNE3), also called Nesprin3, is a spectrin-repeat containing protein with a C-terminal Klarsicht, ANC-1, and Syne Homology (KASH) domain. It is a member of a larger KASH protein family which also includes its paralogs SYNE1 (Nesprin1) and SYNE2 (Nesprin2).^48,49^ These proteins are part of the linker of the nucleoskeleton and cytoskeleton (LINC) complex. SYNE proteins are located at the outer nuclear membrane where they bind cytoskeletal elements. SYNE1 and SYNE2 bind to actin via N-terminal calponin homology domains.^50,51^ SYNE3 lacks a calponin homology domain, and instead recruits crosslinking proteins such as plectin, microtubule actin crosslinking factor 1 (MACF1) and dystonin, which in turn bind to thin, intermediate, and microtubule filament networks.^49,52^ The C-terminal KASH domains of SYNE proteins extend into the perinuclear space, where they bind Sad1p, UNC-84 (SUN) protein homotrimers. These SUN proteins span the inner nuclear membrane and bind the nuclear lamina.^53,54^ While *SYNE3* is not implicated in any Mendelian disease, pathogenic variants in *SYNE1*, which has been shown to play an important role in nuclear positioning and mechanotransduction,^55^ cause Autosomal Recessive Spinocerebellar Ataxia type 8 (SCAR8/ARCA1, OMIM: 610743).^56^ Additionally, mice with homozygous *Sun1*, knock out displayed a cerebellar ataxia phenotype. *Sun1* is a LINC complex protein that binds to SYNE1, SYNE2 and SYNE3 in the peri-nuclear space, highlighting the potential for LINC disruption to cause cerebellar ataxia.^57^

To determine potential functional impacts of the *CLMN*::*SYNE3* chimeric transcript, transcriptomic data from human and mouse brains were interrogated. Publicly available RNA-seq data generated from human brain and cerebellum across developmental and post-natal time points^40^ showed *CLMN* expression increased with age, both generally in the brain, but more markedly in the cerebellum. A similar pattern was observed for *SYNE1*. *SYNE2*, however, had stronger expression during prenatal development. In contrast, *SYNE3* had stronger expression at earlier time points, which reduced with age (Fig. 3D). Single nucleus RNA-seq (snRNA-seq) data from the adult mouse BrainCellData atlas were also analysed.^39^ *Syne1* showed a general neuronal expression pattern, whereas *Syne2* and *Syne3* were not strongly expressed in any brain region. *Clmn* was most strongly expressed in the thalamus, predominantly by thalamic projection neurons, and Purkyně neurons within the cerebellum (Fig. 3E). This Purkyně-restricted cerebellar expression pattern of *Clmn*, along with a lack of *Syne3* expression, was also observed in additional independent adult-mouse brain snRNA-seq data.^58^ Immunostaining of adult mouse brains also showed strong expression of Clmn in cerebellar Purkyně neurons, cerebral cortex, hippocampus, and thalamus.^59,60^ As the *CLMN*::*SYNE3* fusion is expressed from the *CLMN* promoter, these data suggest that the fusion protein would also be most abundant in cerebellar Purkyně neurons. No post-mortem cerebellar tissue was available from affected family members to confirm this hypothesis.

Previous functional studies of recombinant SYNE3 demonstrated that N-terminal epitope tags did not impact the normal function of SYNE3.^61,62^ Thus, we predicted that it was unlikely that the small N-terminal peptide present in the fusion protein would alter the function of SYNE3. Ectopic expression of *SYNE3* results in SYNE3-mediated recruitment of vimentin^62^ and plectin^61^ to the outer nuclear membrane in COS-7 cells.^49^ Additionally, co-expression of SYNE2 and SYNE3 in HaCaT cells led to a reduction in nuclear size.^61^ We generated expression vectors for SYNE3 and the CLMN::SYNE3 fusion, both with N-terminal mEmerald tags, along with an mEmerald empty vector. Transient transfection of *SYNE3* and the *CLMN*::*SYNE3* fusion, hence referred to as SYNE3 constructs, led to recruitment of both vimentin and plectin to the outer nuclear membrane of HeLa and COS-7 cells, which was not seen in cells transfected with the mEmerald empty-vector (Fig. 4A-B, Supplemental Fig. 4). Localisation of SYNE3 and the CLMN::SYNE3 fusion proteins was predominantly perinuclear, compared to the cytoplasmic signal seen in cells transfected with the empty vector (Fig. 4A-B, Supplemental Fig. 4). Further, a statistically significant reduction in nuclear area was observed when comparing HeLa cells transfected with the Empty-Vector to either of the SYNE3 (p=9.5e-11) or CLMN::SYNE3 fusion (p=3.2e-06) constructs (Fig. 4C). These data demonstrated that the CLMN::SYNE3 fusion protein had similar cellular localisation and likely similar interaction with known cytoplasmic binding proteins as wild-type SYNE3.

**Figure 4:**
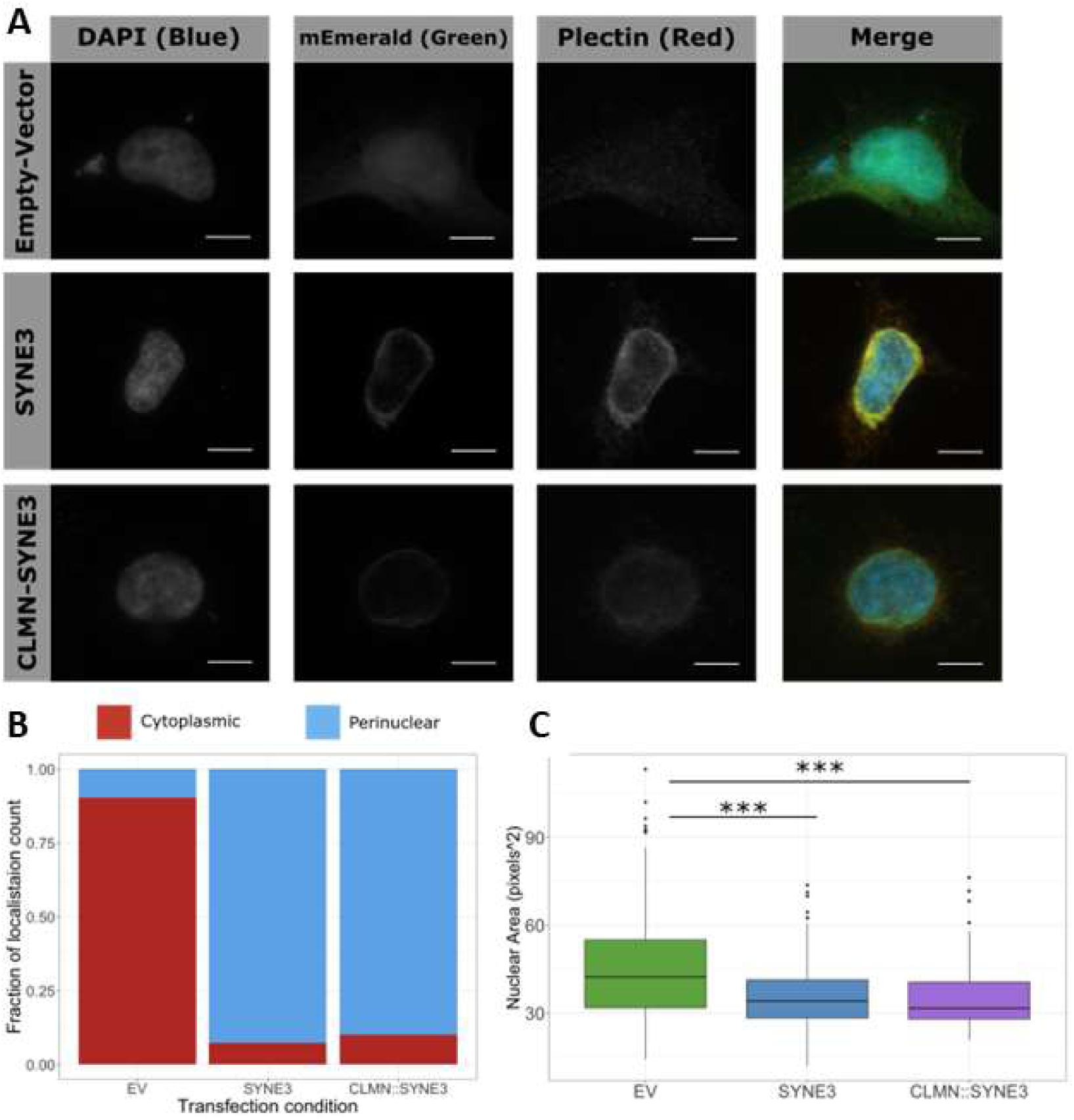
Both SYNE3 and the CLMN::SYNE3 fusion recruit plectin to the nuclear membrane. (A) Representative immunofluorescence images of HeLa cells transiently transfected with mEmerald N-terminally tagged SYNE3, CLMN::SYNE3, or mEmerald empty vector, immunostained with a primary antibody for plectin raised in mouse (Sigma-Aldrich: P9318, RRID: AB_261248, 18.2 μg mL^-1^), and donkey-anti-mouse secondary antibody (Thermo Fisher Scientific: A-31570, RRID: AB_2536180, Alexa-555, 1.33 μg mL^-1^). Individual channels are shown as greyscale images, and merged images shown in colour. Scale bars are 10 µm. Plectin is recruited to the outer nuclear membrane by SYNE3 and the CLMN::SYNE3 fusion, as evident by the rings of co-stained green and red present around the nucleus of these cells, which are absent in the mEmerald empty vector transfected cells. (B) Plot showing localisation of plectin across transfection condition shows predominance of perinuclear staining for HeLa cells transfected with the SYNE3 or CLMN::SYNE3 Fusion expression vectors. Cells counted for each transfection condition: Empty Vector (178), SYNE3 (95), CLMN::SYNE3 Fusion (60). (C) Plot showing distribution of nuclear area in pixels^2^ across transfection conditions reveals a reduction in nuclear size in cells transfected with the SYNE3 or CLMN::SYNE3 expression vectors. A statistically significant reduction in area was seen between both the Empty-Vector and SYNE3 and CLMN::SYNE3 (***=p<0.001, pairwise t-test). Cells counted for nuclear size measurements: Empty Vector (287), SYNE3 (189), CLMN::SYNE3 Fusion (75).

To elucidate the impact of ectopic expression of SYNE3 and the CLMN::SYNE3 fusion proteins in the neuronal context, primary mouse cortical and hippocampal neurons derived from E17.5 mouse embryos were transfected with either the mEmerald empty vector, or the SYNE3 constructs at *in vitro* day 4. In agreement with the observations in HeLa cells, neurons transfected with either of the SYNE3 constructs showed predominantly perinuclear localisation of the recombinant proteins compared to empty vector transfected cells. In some neurons transfected with the SYNE3 constructs, foci were observed extending out into the neuronal appendages (Fig. 5 A-B). We classified nuclear morphology of transfected neurons from the appearance of DAPI staining into three categories: normal, small and round, or fragmented. Neurons transfected with either of the SYNE3 constructs had approximately double the fraction of nuclei that appeared small and round or fragmented, when compared to neurons transfected with the empty vector at both the 24- and 72-hour time points (Fig. 5C). These data suggest that expression of both *SYNE3* and *CLMN*::*SYNE3* in neurons is damaging to their nuclei and likely to negatively impact the long-term survival of these cells.

**Figure 5:**
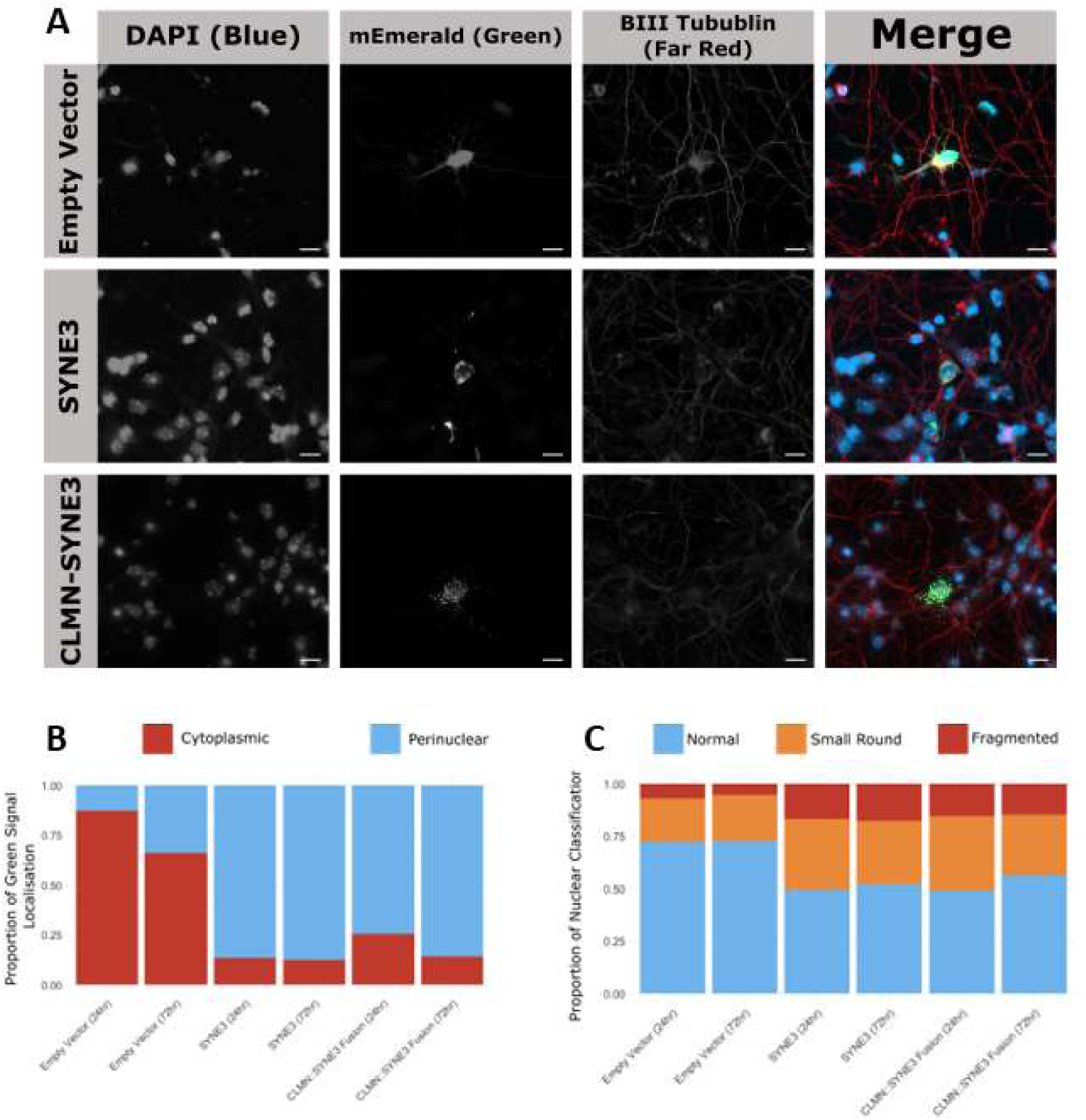
SYNE3 and the CLMN::SYNE3 fusion proteins primarily localise to the nuclear membrane of neurons, with foci forming in axons and dendrites. Representative immunofluorescence images of primary cortical mouse neurons transfected with SYNE3 or CLMN::SYNE3 tagged with mEmerald, or the mEmerald empty vector. Neurons were stained with a primary antibody for β-III Tubulin raised in rabbit (Sigma-Aldrich: T2200, RRID: AB_262133, 2 μg mL^-1^), and donkey-anti-rabbit secondary antibody (Thermo Fisher Scientific: A-31573, RRID: AB_2536183, Alexa-647, 1.33 μg mL^-1^). Individual channels are shown as greyscale images, and merged images shown in colour. Scale bars are 10 µm. (B) Quantitation of recombinant protein localisation in transfected neurons across transfection conditions and time points. Neurons transfected with the SYNE3 or CLMN::SYNE3 expression vectors had predominantly perinuclear localisation at all time points. (C) Plot showing the proportions of transfected neuronal nuclei classifications. Transfection with SYNE3 or the CLMN::SYNE3 leads to an increase in nuclei that appear small and round or fragmented. The number of cells counted for each condition across two replicates were: Empty Vector 24 hours (300) and 72 hours (300), SYNE3 24 hours (239) and 72 hours (234), CLMN::SYNE3 24 hours (300) and 72 hours (247).

## Discussion

We propose that ectopic expression of functional SYNE3 in Purkyně neurons, driven by the *CLMN* promoter, is the likely cause of SCA30. Overexpression of the KASH domains of SYNE proteins can lead to mis-localisation of endogenously expressed SYNEs to the endoplasmic reticulum (ER) and a reduction in mechanical stiffness.^63^ Additionally, the N-terminal spectrin repeats and calponin homology domain of SYNE1 can bind to SYNE3.^61,62^ Thus, the presence of SYNE3 in a cell type where it is not typically expressed could prevent SYNE1 from performing its normal cellular function, disrupting the LINC complex, and mimicking the homozygous loss-of-function of SYNE1 in SCAR8. When the LINC complex is disrupted, transmission of mechanical stimuli from the cytoskeleton to the nucleus and transcriptional response to mechanical stimuli can be disrupted.^64^ For example, myoblasts carrying homozygous *SYNE1* loss of function variants showed an increase in the nuclear localisation of yes-associated protein (YAP), a mechanosensitive transcription factor.^65^ The importance of mechanotransduction pathways in the pathogenesis of neurodegenerative disorders and other forms of ataxia is well known.^66^ Ataxia telangiectasia (A-T) is a multisystemic disorder with a range of symptoms, including loss of Purkyně neurons leading to cerebellar ataxia. A-T is caused by pathogenic variants in Ataxia telangiectasia mutated (*ATM,* OMIM: 208900).^67^ ATM can be expressed in response to oxidative stress and double stranded DNA breaks,^68^ however recent research has demonstrated that *ATM* can also be activated in response to cellular stretching. It was also shown that cells with pathogenic *ATM* variants had an increase in both cellular stiffness and nuclear localisation of YAP.^69^ In the SCA30 family, we detected the *CLMN*::*SYNE3* transcript in LCL which are non-adherent, and less subject to forces acting on the cytoskeleton. Furthermore, *SYNE3* is endogenously expressed in LCLs and not upregulated significantly because of the duplication or the *CLMN*::*SYNE3* fusion which may account for the relatively minor effects on the entire transcriptome we observed. Further functional validation is required to fully elucidate the mechanism of disease in SCA30. Analysis of Purkyně neuron cultures^70^ or cerebellar organoids^71^ generated from patient derived induced pluripotent stem cells (iPSCs) would test the functional impacts of ectopic expression of the *CLMN*::*SYNE3* chimeric transcript, in disease-relevant tissues.

While the importance of chimeric transcripts is well established in the cancer setting,^72,73^ their role in Mendelian disease is less well characterised. Mendelian disorders currently have an approximate genetic diagnostic yield of around 30-50%,^74,75^ however this number is increasing in a disease-dependent manner. Previously undetected disease-causing SVs and CNVs have been identified through the application of PacBio^76^ and ONT^77^ long-read GS and Bionano optical genome mapping.^78^ Further, the application of methylation signatures^79^ and RNA-seq data^36^ have allowed the reclassification of variants of unknown significance. Despite these advances, there are still many genetic causes of disease yet to be identified. One potential source of these missing diagnoses are SVs that give rise to chimeric transcripts. Fusion transcripts arising from deletions and reciprocal translocations have been identified in a range of inherited disorders, however, in these cases the suggested disease mechanism is haploinsufficiency of the affected genes.^80–83^ In a study that analysed a cohort of 112 individuals with 119 tandem duplications referred for diagnostic cytogenetic testing, a total of 21 chimeric RNA species were predicted.^84^ Six of these produced in-frame fusions that were hypothesised to play a role in disease.^84^ There has been an influx of bioinformatic tools designed to detect chimeric transcripts from long and short read RNA-seq data,^85,86^ however RNA sequencing is not performed routinely in the genomic diagnostic setting, hampering detection of chimeric RNAs and limiting full interpretation of the consequences of CNV and SV on the transcriptome.

Chimeric RNA species could also be an appealing target for treatments. The unique splice junctions formed in chimeric RNA species could be targeted by antisense oligonucleotides (ASOs). These short nucleotide sequences are designed to specifically reduce mutant protein levels by targeting transcripts for degradation.^87^ ASOs have been used in clinical trials aimed at treating other neurodegenerative diseases such as amyotrophic lateral sclerosis^88^ and Huntington disease.^89,90^ ASOs show high efficacy when targeting transcripts that are not usually present in healthy cells. One such example is the pseudo-exon introduced into *COL6A1* transcripts in the presence of a deep intronic variant that causes Collagen VI-related congenital muscular dystrophies.^91^ ASOs have been shown to effectively target this mutant transcript, and restore wild-type *COL6A1* transcripts.^91^ As such, the *CLMN*::*SYNE3* chimeric transcript, which is normally absent in human cells, could also be a potential target for degradation with ASOs. Through extensive genetic and transcriptomic analysis, we identified a tandem duplication within 14q32.13 that leads to the production of a novel splice-mediated chimeric transcript between *CLMN* and *SYNE3*, which we suggest is the cause of SCA30. A limitation of this study is the confinement of this variant to a single, large family where it is fully explanatory of the phenotype. Of note, founder events are a common occurrence in SCAs where the late age onset phenotype does not impose strong selection. The implication of the *CLMN::SYNE3* chimeric transcript in SCA30 goes beyond resolving a single family by highlighting an underappreciated molecular consequence of CNV and SV that has high potential for targeted treatment with ASO technology.

## Data Availability

GS and RNA-seq data generated in this study is available via request to the corresponding author and is subject to approval for use by the Womens and Childrens Health Network human research ethics committee.
Mouse single nuclei data is available from https://docs.braincelldata.org/downloads/index.html
Human developmental bulk RNA seq data from EvoDevo is available from https://www.ebi.ac.uk/biostudies/arrayexpress/studies/E-MTAB-6814

https://www.ebi.ac.uk/biostudies/arrayexpress/studies/E-MTAB-6814

https://neuromuscular.wustl.edu/ataxia/domatax.html

## Acknowledgements

We wish to thank the family involved in this study.

## Funding

This work was supported by funding from the following sources: University of Adelaide, Adelaide Medical School (MAC), National Health and Medical Research Council Synergy grant 2010562 (JG), Centre for Research Excellence 1117394 and Senior Research Fellowship 1155224 (JG), Australian Government Genomics Health Futures Mission grants MRFF2024989 (PJLockhart) and MRFF2007681 (GM, NGL), Channel 7 Children’s Research Foundation (CAS), Women’s and Children’s Hospital Research Foundation (CAS), the Independent Research Institute Infrastructure Support Scheme and the Victorian State Government Operational Infrastructure Program (PJLockhart).

## Competing interests

The authors declare that they have no competing interests.

## Author contributions

Conceptualization: JG, PJLockhart

Data curation: TML, RC, MAC

Formal analysis: TML, MAC, AEG, KLF, MA, HR, GM, MB

Funding acquisition: JG, MAC, GR, PJLockhart, CAS

Investigation: TML, RAW, KLF, GM, AEG, MA, KL, LS, NMN, CAS, MBD, PJLamont, CA

Methodology: TML, MAC, NMN

Project administration: JG, MAC

Resources: RAW, EH, MBD, MA, CAS, PJLamont, CA, PJLockhart,

Software: TML

Supervision: JG, MAC, GR, NGL, MB, PJLockhart

Validation: TML, MAC

Visualization: TML

Writing – original draft: TML

Writing – review & editing: All authors

## Supplementary material

Supplementary material is provided at the end of this document.

---

**Supplemental Figure 1:**
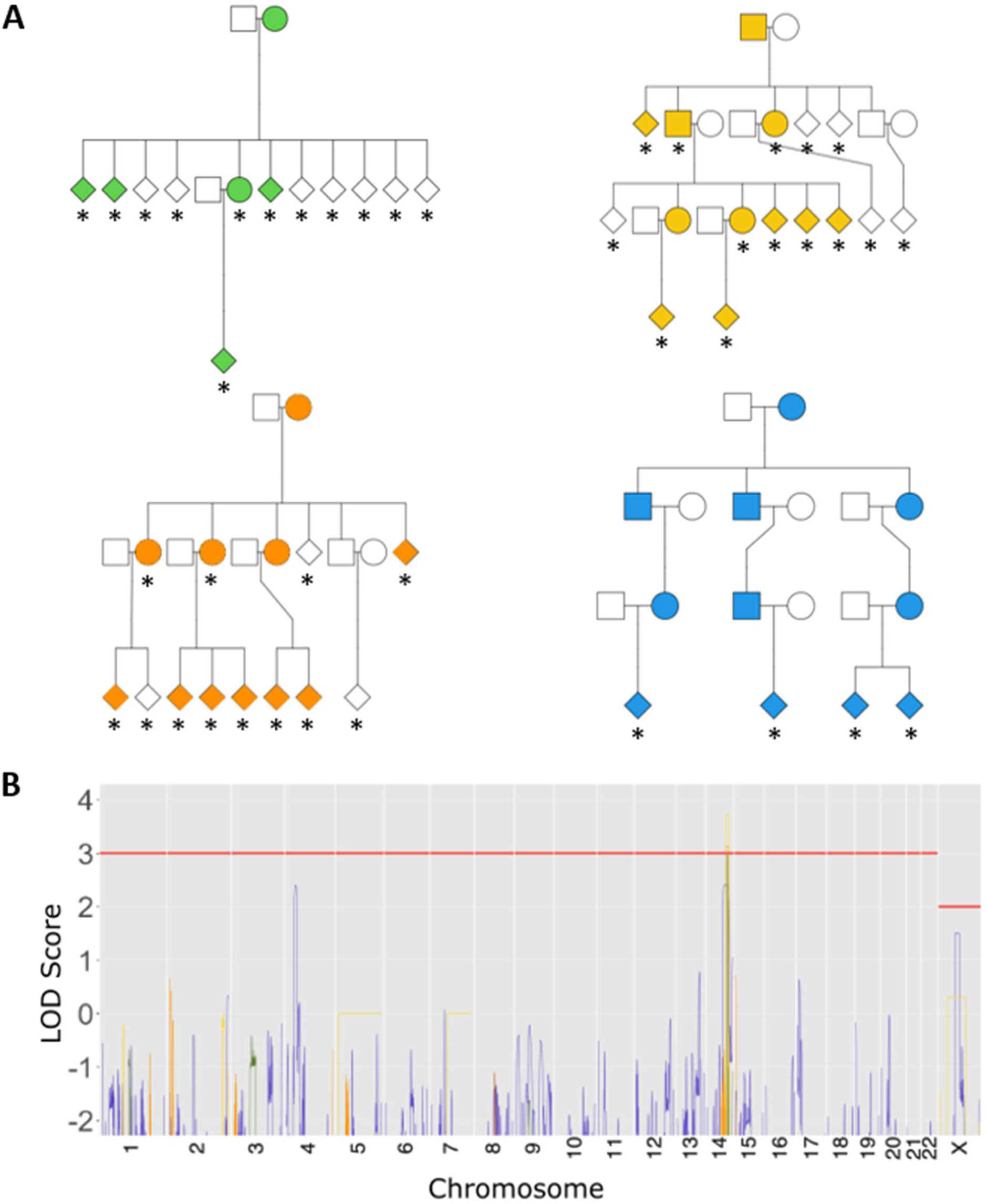
Non-overlapping sub-pedigrees used to complete genome-wide parametric linkage analysis show a shared linkage signal on chromosome 14. (A) Pedigree showing breakdown of non-overlapping sub-pedigrees used for linkage analysis. Males are marked with squares and females with circles. Affected individuals are marked with black shapes and unaffected individuals are marked with white shapes. Individuals with genotyping data included in the linkage analysis are marked with an *. (B) Plot shows the LOD scores generated from genome-wide parametric linkage analysis for all SNP markers based on their position in the human genome from chromosome 1-22 and X for each of the four sub-pedigrees. The red horizontal lines LOD denote significance thresholds of 3 for autosomes and 2 for the X chromosome.

**Supplemental Figure 2:**
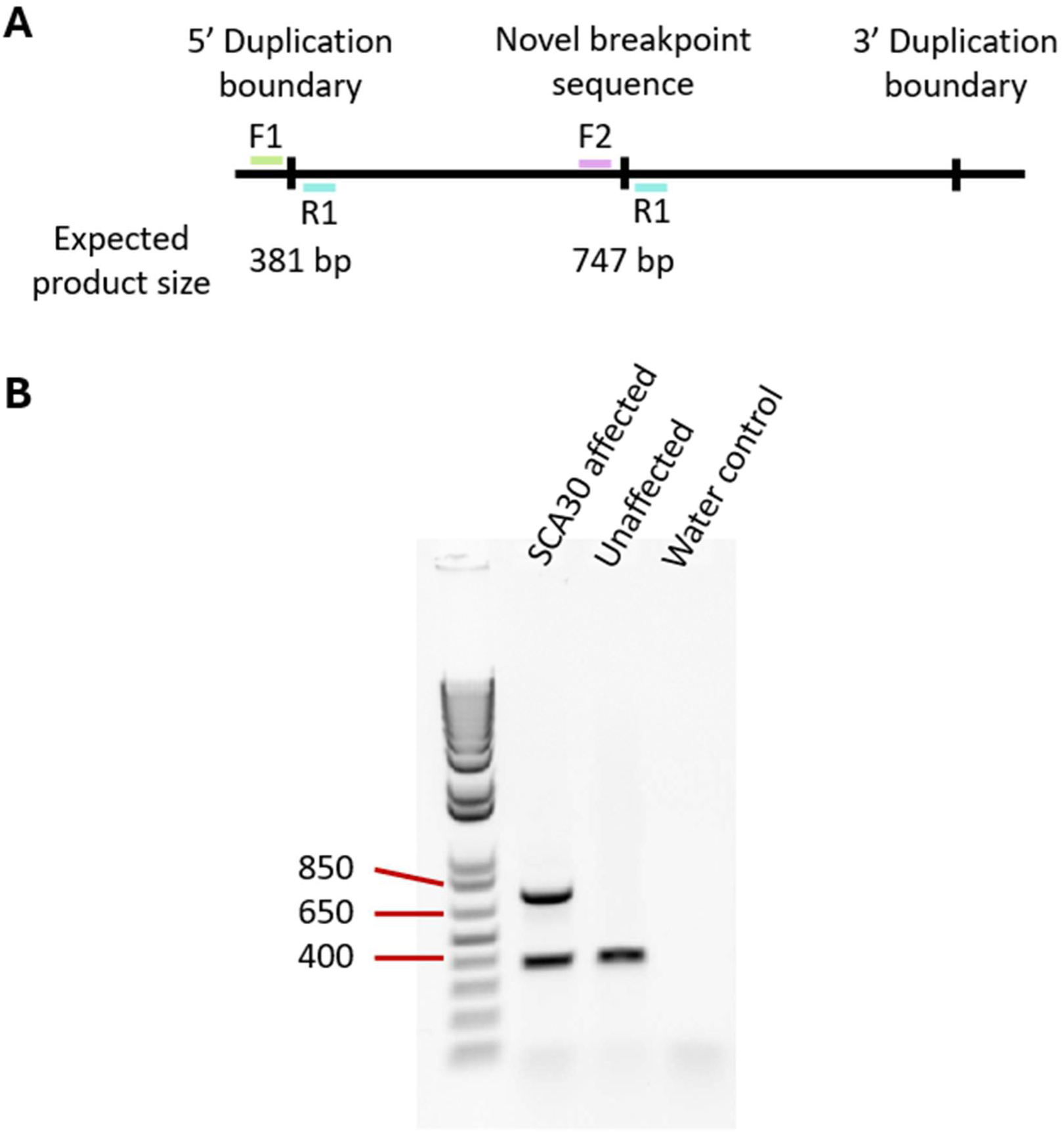
Diagrammatic representation of duplex genomic PCR used to segregate and screen for the SCA30 duplication. (A) Diagram shows the expected fragment size of the 5’ duplication boundary PCR, which should work for all individuals, and the PCR that amplifies the unique genomic sequence present only in those with the SCA30 duplication. (B) Example agarose gel showing expected banding pattern for duplex PCR for an individual with SCA30 (left), an unaffected individual (middle) and the negative water control (right).

**Supplemental Figure 3:**
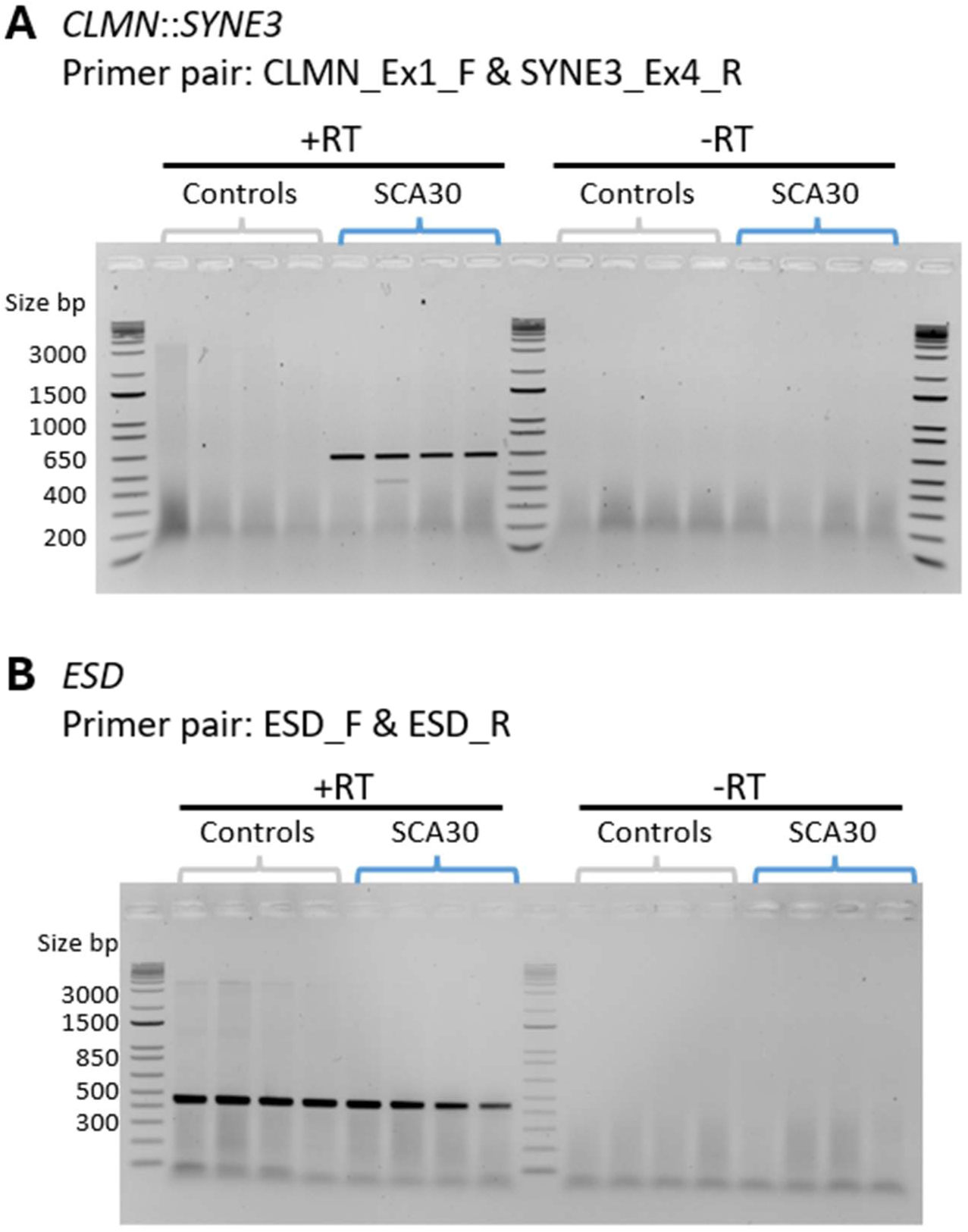
Detection of the *CLMN*::*SYNE3* transcript by RT-PCR. PCR products for (A) *CLMN*::*SYNE3* using primers CLMN_Ex1_F and SYNE3_Ex4_R (Supplemental Table S1) and (B) *ESD* derived from cDNA prepared from total RNA extracted from LCL of four female individuals who are not affected by SCA and unrelated to the SCA30 family compared to four affected individuals from the SCA30 family (V.56, V.61, VI.16 and V.82). PCR products were only amplified from samples where the reverse transcriptase was included in the reaction to produce cDNA (+RT) and not where it was excluded (-RT) indicating that these products are derived from mRNA transcripts spanning multiple exons. The *CLMN*::*SYNE3* product corresponds with a predicted 683 bp spanning *CLMN* Exon 1 to *SYNE3* Exon 4 was only observed in affected individuals from the SCA30 family. The 453 bp amplicon from the ubiquitously expressed *ESD* gene indicates cDNA template is present in all +RT samples. Molecular weight markers are 1kb Plus ladder (Thermo Fisher Scientific cat. # 10787018).

**Supplemental Figure 4:**
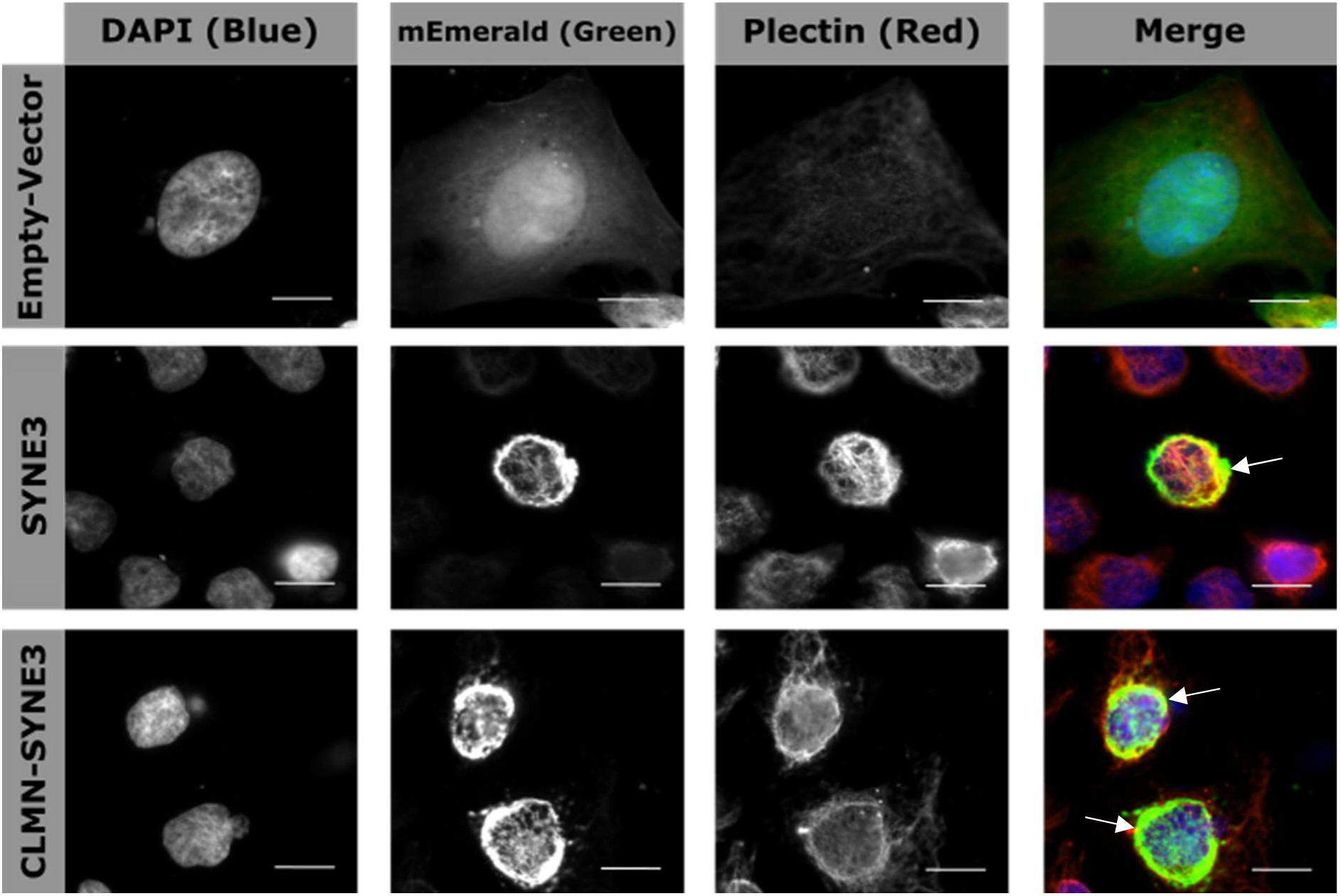
SYNE3 and CLMN::SYNE3 fusion proteins recruit plectin to the nuclear membrane of HeLa cells. Images taken at 63x magnification of HeLa cells transiently transfected with either the mEmerald-empty-vector, mEmerald-SYNE3, or mEmerald-CLMN::SYNE3 fusion. Cells were incubated with primary antibody for vimentin raised in mouse (Thermo Fisher Scientific: T2200, 2 μg ml^-1^), and donkey-anti-mouse secondary antibody (Invitrogen: A3157, Alexa-555, 1.33 μg ml^-1^). Individual channel images are shown in grey scale with the merged image shown in colour. Scale bar is 10 µm. Recruitment of vimentin to the nuclear membrane shown by co-staining of green and red signal around the nucleus of cells transfected with SYNE3 and CLMN::SYNE3, but not in those transfected with the empty vector.

**Table S1.**
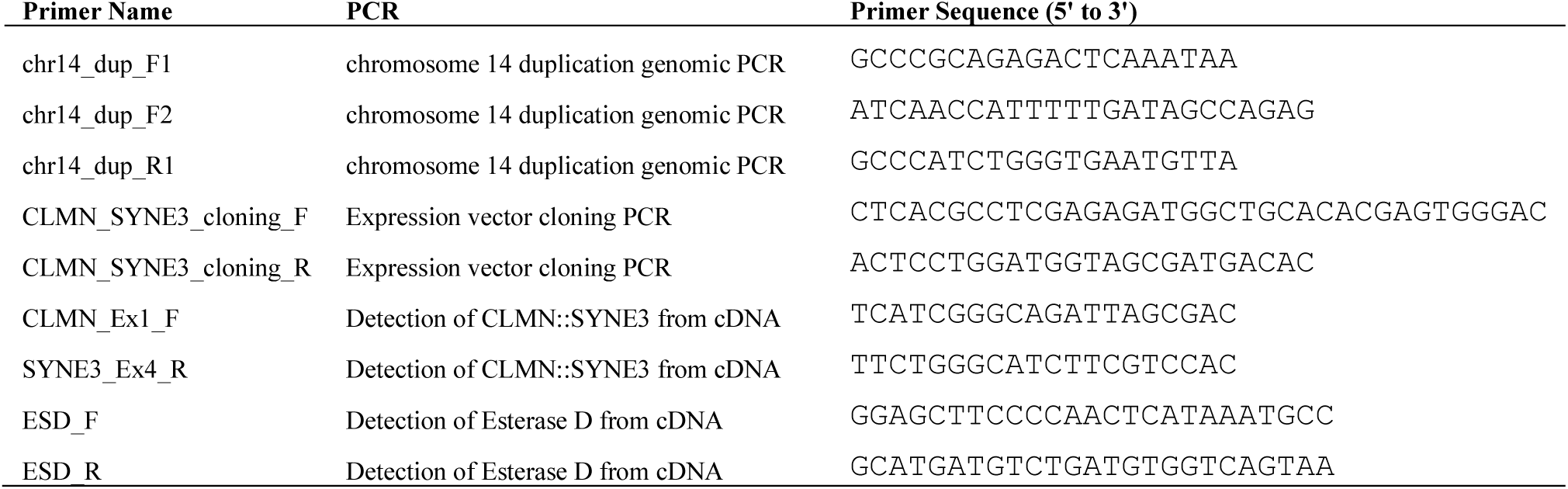
Primer sequences.

**Table S2.** Primary antibodies.

| **Antibody** | **Source Species** | **Company** | **Catalogue** | **Dilution** |
| --- | --- | --- | --- | --- |
| Anti-Plectin | Mouse | Sigma | P9318 | 18.2 µg ml<sup>-1</sup> |
| Anti-Vimentin | Mouse | Thermo Fisher Scientific | MA5-11883 | 1 µg ml<sup>-1</sup> |
| Anti-β-3 Tubulin | Rabbit | Thermo Fisher Scientific | T2200 | 2 µg ml<sup>-1</sup> |

**Table S3.** Secondary antibodies.

| **Target Species** | **Source Species** | **Fluorophore** | **Company** | **Catalogue** | **Dilution** |
| --- | --- | --- | --- | --- | --- |
| Rabbit | Donkey | Alexa-647 (Far-red) | Invitrogen | A31573 | 1.33 $\mu\text{g ml}^{-1}$ |
| Mouse | Donkey | Alexa-555 (Red) | Invitrogen | A31570 | 1.33 $\mu\text{g ml}^{-1}$ |

**Table S4.** Shared rare and private variants identified within the chromosome 14 linkage interval.

| **Chr** | **Start** | **Ref** | **Alt** | **dbSNP** | **Functional Element** | **MAF<sup>a</sup>** | **CADD<sup>b</sup>** | **ReMM<sup>c</sup>** |
| --- | --- | --- | --- | --- | --- | --- | --- | --- |
| 14 | 94353020 | A | T | NA | NA | 3.19E-05 | 2.534 | 0 |
| 14 | 95545106 | G | A | rs99346944 | Enhancer (Downstream - GLRX5) | 6.37E-05 | 0.066 | 0.0033 |

<sup>a</sup> MAF; Minor allele frequency in gnomAD v2.1.1.
<sup>b</sup> CADD; Combined annotation dependent depletion.
<sup>c</sup> ReMM; Regulatory Mendelian Mutation

**Table S5.** Shared structural variants within linkage interval identified from Oxford Nanopore genome sequencing.

| **Chr** | **Start** | **End** | **Type** | **Size (bp)** | **Affected Genes** | **ACMG Classification (AnnotSV)** |
| --- | --- | --- | --- | --- | --- | --- |
| 14 | 95239046 | 95570197 | Duplication<br>(Tandem) | 331151 | *CLMN, SNHG10, SYNE3, GLRX5* | 3 |

**Table S6.** Differentially expressed genes identified from SCA30 RNA-seq data.

| **Gene symbol** | **logCPM<sup>a</sup>** | **logFC<sup>b</sup>** | **P-value** | **FDR<sup>c</sup>** |
| --- | --- | --- | --- | --- |
| *DDX39B* | 6.905765717 | 9.739256335 | 3.02E-39 | 2.36E-35 |
| *SMG1P1* | 5.211046836 | -1.642887678 | 2.39E-08 | 3.41E-05 |
| *KCNQ2* | -1.673807259 | -3.140043842 | 1.00E-07 | 0.00013042 |
| *DPY19L1P1* | -0.83523161 | -2.416786356 | 2.37E-07 | 0.00028356 |
| *TMEM191B* | 0.878469924 | -2.297060639 | 2.54E-07 | 0.00028356 |
| *PFKFB4* | 6.759937365 | -0.982727822 | 5.26E-07 | 0.00054842 |
| *FMNL2* | 3.132502246 | -1.390535587 | 1.28E-06 | 0.00118213 |
| *ARHGAP19-SLIT1* | -0.8749156 | -4.25957751 | 1.37E-06 | 0.00118955 |
| *PPFIA4* | 5.364969149 | -1.037831394 | 1.45E-06 | 0.00119327 |
| *CLEC18C* | -2.146375624 | 4.445207788 | 4.49E-06 | 0.00334592 |
| *GLRX5* | 5.188757691 | 0.988915272 | 5.55E-06 | 0.0039442 |
| *SAXO2* | 1.167398062 | 4.386745477 | 7.24E-06 | 0.0049252 |
| *FUT11* | 5.526616342 | -1.022044993 | 7.67E-06 | 0.00499612 |
| *HILPDA* | 4.849747196 | -1.626258771 | 1.22E-05 | 0.00739037 |
| *TCAF2* | 5.065585701 | -0.887269691 | 1.23E-05 | 0.00739037 |
| *RHOBTB1* | -1.333746352 | -1.872078986 | 1.92E-05 | 0.01113421 |
| *SLC16A3* | 7.043944444 | -0.759534043 | 2.17E-05 | 0.01213893 |
| *ZNF460* | 3.097052524 | -1.273013288 | 2.30E-05 | 0.01230616 |
| *ABHD16A* | 2.6899475 | 2.384979895 | 2.36E-05 | 0.01230616 |
| *SH3D21* | 4.399199183 | -0.758252766 | 2.74E-05 | 0.01383766 |
| *PFKFB3* | 5.655242025 | -0.828940519 | 2.96E-05 | 0.01444761 |
| *BMP4* | -0.435204819 | 3.104442075 | 4.20E-05 | 0.0193136 |
| *MXI1* | 4.735406244 | -0.873743588 | 5.22E-05 | 0.02334006 |
| *FGF11* | 5.081259673 | -0.789472794 | 6.13E-05 | 0.02661999 |
| *KMT2C* | 7.548225252 | -0.865974072 | 6.33E-05 | 0.02674707 |
| *THOC3* | 5.684305825 | -0.687038477 | 6.88E-05 | 0.0283366 |
| *PI4KAP1* | 4.549032885 | -1.655369299 | 8.84E-05 | 0.03457703 |
| *LINC03040* | 4.938458689 | -1.026198707 | 0.000100562 | 0.03836569 |
| *TCL1A* | 6.774103537 | -0.776328578 | 0.000105495 | 0.03928927 |
| *KDM6B* | 6.482568663 | -0.691333944 | 0.000126772 | 0.04406599 |
| *KDM3A* | 6.990386777 | -0.659007235 | 0.000133334 | 0.04470509 |
| *IGHV5-10-1* | 2.978052228 | -9.185938865 | 0.000134327 | 0.04470509 |

<sup>a</sup> log 2 transformed transcript counts per million mapped reads (CPM) summarized to the level of the gene
<sup>b</sup> log 2 transformed fold change in gene expression. A negative value implies the gene is down-regulated in SCA30 samples and a positive value implies the gene is up-regulated in SCA30 samples.
<sup>c</sup> False discovery rate

**Table S7.** Pathways identified through gene ontology enrichment analysis with ShinyGO.

| Enrichment FDR | nGenes | Pathway Genes | Fold Enrichment | Pathway | Genes | Pathway Database |
| --- | --- | --- | --- | --- | --- | --- |
| 0.002937648 | 2 | 4 | 254.962963 | GO:0003873 6-phosphofructo-2-kinase activity | *PFKFB4, PFKFB3* | GO Molecular Function |
| 0.002937648 | 2 | 5 | 203.9703704 | GO:0004331 fructose-2,6-bisphosphate 2-phosphatase activity | *PFKFB4, PFKFB3* | GO Molecular Function |
| 0.002937648 | 2 | 11 | 169.9753086 | GO:0050308 sugar-phosphatase activity | *PFKFB4, PFKFB3* | GO Molecular Function |
| 0.002937648 | 2 | 7 | 145.6931217 | GO:0008443 phosphofructokinase activity | *PFKFB4, PFKFB3* | GO Molecular Function |
| 0.002937648 | 2 | 12 | 145.6931217 | GO:0019203 carbohydrate phosphatase activity | *PFKFB4, PFKFB3* | GO Molecular Function |
| 0.016180306 | 2 | 17 | 92.71380471 | GO:0000346 transcription export complex | *THOC3, DDX39B* | GO Cellular Component |
| 0.016180306 | 2 | 15 | 84.98765432 | GO:0044666 MLL3/4 complex | *KMT2C, KDM6B* | GO Cellular Component |
| 0.019646639 | 2 | 21 | 53.67641326 | GO:0019200 carbohydrate kinase activity | *PFKFB4, PFKFB3* | GO Molecular Function |
| 0.029954356 | 2 | 32 | 37.77229081 | GO:0032452 histone demethylase activity | *KDM3A, KDM6B* | GO Molecular Function |
| 0.029954356 | 2 | 32 | 37.77229081 | GO:0140457 protein demethylase activity | *KDM3A, KDM6B* | GO Molecular Function |
| 0.040116428 | 2 | 5 | 203.9703704 | GO:0006003 fructose 2,6-bisphosphate metabolic proc. | *PFKFB4, PFKFB3* | GO Biological Process |
| 0.040116428 | 2 | 13 | 145.6931217 | GO:0046784 viral mRNA export from host cell nucleus | *THOC3, DDX39B* | GO Biological Process |
| 0.040116428 | 2 | 14 | 127.4814815 | GO:0044417 translocation of molecules into host | *THOC3, DDX39B* | GO Biological Process |
| 0.047273662 | 2 | 42 | 28.32921811 | GO:0032451 demethylase activity | *KDM3A, KDM6B* | GO Molecular Function |

## Notes

### Competing Interest Statement

The authors have declared no competing interest.

### Author Declarations

The Human Research Ethics Committee of the Womens and Childrens Health Network gave ethical approval for this work (HREC2361/3/2026). The Human Research Ethics Committee of The Royal Childrens Hospital gave ethical approval for this work (HREC #28097). The Medical Animal Ethics Committee of the University of Adelaide (now Adelaide University) gave ethical approval for this work (M-2023-017).

